# Assay-related Differences in SuPAR Levels: Implications for Measurement and Data Interpretation

**DOI:** 10.1101/2021.06.23.21259148

**Authors:** Salim S. Hayek, Laura M. Raffield, Yan Gao, Gunnar Engstrom, Arshed A. Quyyumi, Alexander P. Reiner, Jochen Reiser

**Affiliations:** University of Michigan, Department of Medicine, Division of Cardiology, Ann Arbor MI; Department of Genetics, University of North Carolina, Chapel Hill, North Carolina; Jackson Heart Study, University of Mississippi Medical Center, Jackson, Mississipi; Department of Clinical Sciences, Lund University, Malmö, Sweden; Division of Cardiology, Emory University School of Medicine, Atlanta, GA; Department of Epidemiology, University of Washington, Seattle, Washington; Department of Medicine, Rush University Medical Center, Chicago, IL, USA

**Keywords:** SOMAScan, Olink, Quantikine, suPARnostic, Virogates, R&D, soluble urokinase plasminogen activator receptor

## Abstract

With the growing interest in studying the role of soluble urokinase plasminogen activator receptor (suPAR) in kidney and other diseases, levels are now being measured in major cohorts across the world through various methods, including immunoassays and through proteomics. Reported suPAR values have however varied dramatically depending on which assay is used. Comparing suPAR assays and their ability to discriminate risk is crucial for the Nephrology and overall research community to guide assay choice and inform interpretation of findings. To that end we measured suPAR in 3 different major cohorts (Malmo Diet and Cancer Study, n=4637; Jackson Heart Study, n=1905; and the Emory Cardiovascular Biobank, n=487) using two different methods in each cohort, encompassing the most commonly used approaches (immunoassays and proteomics). We find dramatic differences between assays; with notably poor correlations between immunoassays and proteomic approaches (r=0.2-0.5). Proteomics-based suPAR measures very poor discriminatory ability for cardiovascular and incident kidney disease. Amongst the immunoassays, the suPARnostic assay reported the highest values and had the strongest discriminatory ability.

---

Soluble urokinase plasminogen activator receptor (suPAR) is an immune-derived glycoprotein implicated in kidney disease and associated with various clinical outcomes(1-3). The rising interest in exploring suPAR’s role in kidney and cardiovascular disease has led to its measurement in various cohorts(1-3). Different assays have been used to measure suPAR levels: enzyme-linked immunosorbent assays (ELISA) include the Human uPAR Quantikine ELISA (R&D Systems, Minneapolis, MN), and the suPARnostic ELISA (Virogates, Copenhagen, Denmark); and proteomics platforms such as the aptamer-based assay SomaLogic SOMAscan (SomaLogic, Boulder, CO) and the proximity extension assay Olink Explore (Olink, Uppsala, Sweden). SuPAR values and associations with clinical outcomes have differed greatly according to the assay used(4), warranting an exploration of assay-related differences in suPAR levels. To that end, we leveraged cohorts in which suPAR was measured using two different approaches, and examined the correlation between assays, the association between levels and clinical characteristics, and their risk discrimination ability for relevant outcomes.

Data on plasma suPAR measurement, clinical characteristics and outcomes were obtained from the following cohorts: a subset (n=4637) of the Malmö Diet and Cancer Study (MDSC) which is a Swedish population-based cohort in whom suPAR was measured using the suPARnostic ELISA and Olink CVD-I panel(2); a subset (n=1492) of the Jackson Heart Study (JHS) which recruited African American participants from Jackson, Mississippi in whom suPAR was measured using the Human uPAR Quantikine ELISA and SOMAscan(3); and lastly, a subset (n=487) of the Emory Cardiovascular Biobank (EmCAB) which enrolled patients with undergoing coronary catheterization, in whom suPAR was measured with the suPARnostic and Human uPAR Quantikine ELISAs(1).

In JHS, SOMAscan values are natural log-transformed, standardized to a mean of 0 and an SD of 1 within each of three batches, and then inverse-normalized across batches. In MDSC, Olink’s normalized protein expression units are log-based 2 transformed. We used Spearman-Rank to report the pairwise correlation between assays. We report the association between clinical characteristics and suPAR measures for each assay separately using linear regression, with suPAR as the dependent variable and clinical characteristics as independent covariates. Lastly, we computed Harrell’s C-statistic as a measure of risk discrimination for each suPAR assay for the following outcomes: all-cause mortality, cardiovascular mortality, and incident chronic kidney disease (defined as a decrease in creatinine-derived eGFR to below 60 ml/min/1.73 m^2^). Analyses were performed using SPSS 24 (IBM, NY, USA) and R (R Core Team, 2014).

We found extensive variation in the correlation between suPAR assays (Table). Measures using the proteomics platforms correlated poorly with the ELISAs, with a correlation of 0.285 between the Human uPAR Quantikine and SOMAScan, and 0.586 between the suPARnostic and and Olink. The correlation between the suPARnostic and Human uPAR Quantikine assay was relatively better at 0.753, with values obtained using the suPARnostic assay on average 50% higher than the Quantikine-derived values, and with significant variability (standard deviation of 50%). Associations with relevant clinical characteristics and their directionality were mostly consistent across measures, except for diabetes mellitus which was not associated with SOMAScan-derived suPAR levels in JHS but was associated with Quantikine ELISA measures in the same individuals (Table). Risk discrimination as quantified using Harrell’s C-statistic for all-cause death, cardiovascular death and incident chronic kidney disease differed between suPAR assays, with both proteomics platforms performing poorly compared to the ELISAs, and suPARnostic outperforming Human uPAR Quantikine (Table).

This study highlights major discrepancies between suPAR measurements across commonly used assays, with poor correlations and differences in associations with outcomes. These findings warrant caution in deriving conclusions related to suPAR measures obtained using proteomics platforms. Proteomics platforms, while useful for discovery, have limitations including cross-reactivity, lack of specificity, protein complexes and single nucleotide polymorphisms altering aptamer/antibody affinities, amongst others which may be more relevant for some assayed proteins such as suPAR(5). Most importantly, proteomics-derived suPAR measures have not been previously cross-validated with ELISAs. We also found significant differences in the ELISA measurements, consistent with a prior report by Winnicki et al. in which suPAR levels using the suPARnostic assay were higher and outperformed the Human Quantikine uPAR ELISA in differentiating between patients with and without focal segmental glomerulosclerosis(4). SuPAR exists in circulation in different forms originating from splice variants and proteolytic processing with varying levels of glycosylation and distinct biological activity. The suPARnostic ELISA consists of two monoclonal capture antibodies; one targeting the D^III^ subunit, and the other the D^II^ subunit, thus capturing full-length suPAR (D^I^D^II^D^II^) and the D^II^D^III^ fragment, but not D^I^. The Human uPAR Quantikine assay uses a monoclonal capture antibody and polyclonal detection antibodies. The discrepancy between the ELISAs may relate to their differing ability in detecting the various suPAR forms. The suPARnostic assay’s ability to detect both full length and cleaved suPAR forms may explain its higher levels and overall better risk discrimination making it the preferred assay for suPAR to determine kidney and cardiovascular outcome measures.

## Data Availability

Data for these studies can be obtained through ancillary proposals directed to the selected cohorts.

## Acknowledgements

The Jackson Heart Study (JHS) is supported and conducted in collaboration with Jackson State University (HHSN268201800013I), Tougaloo College (HHSN268201800014I), the Mississippi State Department of Health (HHSN268201800015I) and the University of Mississippi Medical Center (HHSN268201800010I, HHSN268201800011I and HHSN268201800012I) contracts from the National Heart, Lung, and Blood Institute (NHLBI) and the National Institute on Minority Health and Health Disparities (NIMHD). The authors also wish to thank the staffs and participants of the JHS. The views expressed in this manuscript are those of the authors and do not necessarily represent the views of the National Heart, Lung, and Blood Institute; the National Institutes of Health; or the U.S. Department of Health and Human Services.

---

**Table.**
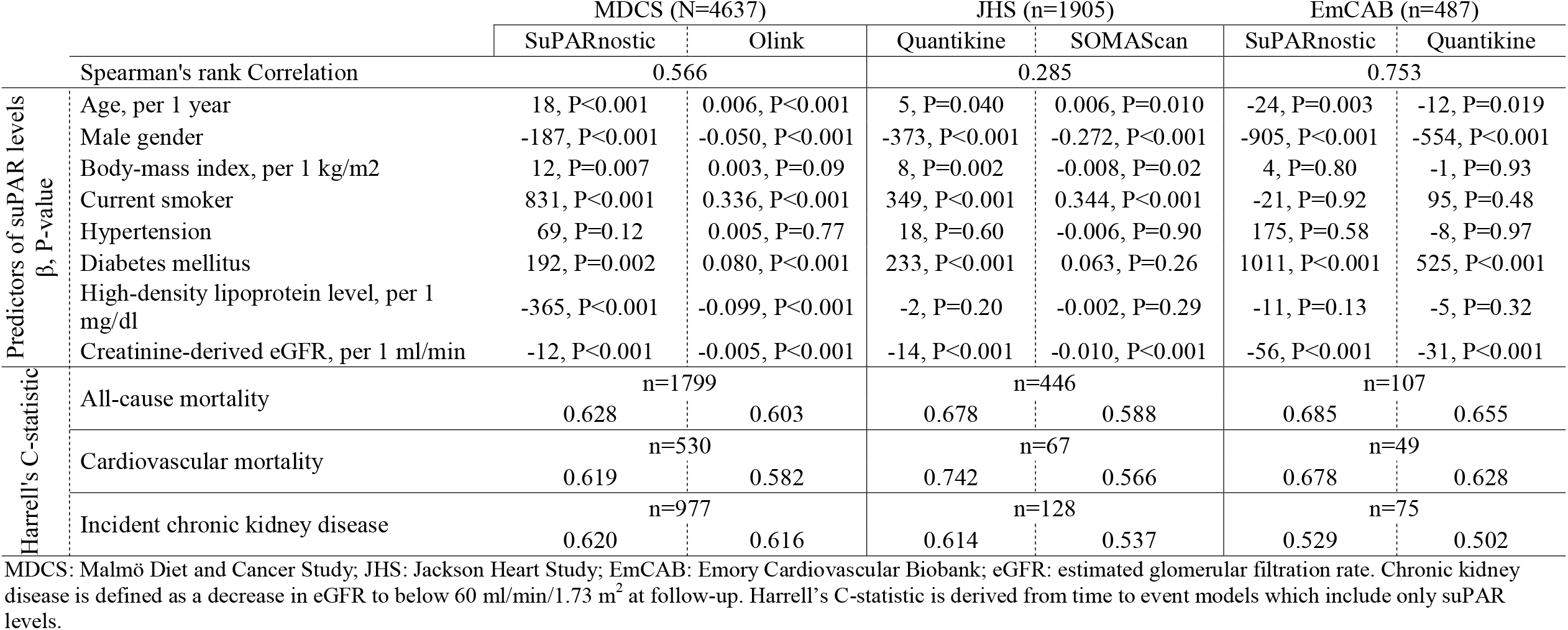
Correlation, associations with clinical characteristics, and C-statistics stratified by cohort and assay.

